# Detecting Alzheimer Disease in EEG Data with Machine Learning and the Graph Discrete Fourier Transform

**DOI:** 10.1101/2023.11.01.23297940

**Authors:** Xavier S. Mootoo, Alice Fours, Chinthaka Dinesh, Mohammad Ashkani, Adam Kiss, Mateusz Faltyn

## Abstract

Alzheimer Disease (AD) poses a significant and growing public health challenge worldwide. Early and accurate diagnosis is crucial for effective intervention and care. In recent years, there has been a surge of interest in leveraging Electroencephalography (EEG) to improve the detection of AD. This paper focuses on the application of Graph Signal Processing (GSP) techniques using the Graph Discrete Fourier Transform (GDFT) to analyze EEG recordings for the detection of AD, by employing several machine learning (ML) and deep learning (DL) models. We evaluate our models on publicly available EEG data containing 88 patients categorized into three groups: AD, Frontotemporal Dementia (FTD), and Healthy Controls (HC). Binary classification of dementia versus HC reached a top accuracy of 85% (SVM), while multiclass classification of AD, FTD, and HC attained a top accuracy of 44% (Naive Bayes). We provide novel GSP methodology for detecting AD, and form a framework for further experimentation to investigate GSP in the context of other neurodegenerative diseases across multiple data modalities, such as neuroimaging data in Major Depressive Disorder, Epilepsy, and Parkinson disease.

## 1 Introduction

Alzheimer Disease (AD) is a progressive and irreversible neurodegenerative disorder characterized by memory loss, impaired problem-solving, reasoning, language difficulties, and physical decline (Scheltens et al., 2016); (Porsteinsson et al., 2021). Advancing age is the predominant risk factor for AD due to the accumulation of abnormal proteins like A*β* plaques and tau tangles, along with reduced neuronal repair capacity during the aging process (Lesné et al., 2006); (Binder et al., 2005); (Knopman et al., 2021). Genetic predisposition and co-morbidities further increase risk for this multi-factorial disease (Rabinovici, 2019); (Lindsay et al., 2002); (Silva et al., 2019).

AD and Frontotemporal Dementia (FTD) represent two distinct yet complex disorders with overlapping clinical features (Götz & Ittner, 2008); (Lindau et al., 2000); (Pachana et al., 1996). FTD is another neurodegenerative disorder affecting cognitive functions. Unlike AD, which predominantly impacts the hippocampal and cerebral cortex regions, FTD is localized to the frontal and temporal lobes of the brain that govern behavior, personality, and language (Olney et al., 2017); (Levy et al., 1996). Subtypes of FTD manifest with specialized symptoms, including behavioral and language variants (Olney et al., 2017); (Pasquier et al., 2004). Given their symptomatic overlaps, distinguishing between AD and FTD in a clinical setting can be challenging (Warren et al., 2013); (Ritter et al., 2017); Rosen et al. (2002).

AD constitutes a significant majority of dementia cases, contributing to approximately 60-70% of diagnoses (Giridharan et al., 2022); (van der Flier & Scheltens, 2005). The World Health Organization (WHO) reported that, in 2020, a total of 50 million individuals lived with dementia globally, with projections suggesting a rise to 152 million by 2050. (Nichols et al., 2022); (Houmani et al., 2018); (Giridharan et al., 2022). Dementia-related disorders are diagnosed upon the emergence of symptoms through neurological assessments and managed through lifestyle modifications aimed at attenuating cognitive decline (Yiannopoulou & Papageorgiou, 2020); (Wahl et al., 2019); (Dominguez et al., 2021). With an aging demographic, there is a pressing need for the implementation of diagnostic tools capable of detecting the onset of these diseases (Knopman et al., 2021).

AD shows distinct alterations in the synchronicity and complexity of brain waves compared with normal brain and other types of dementia (Zande et al., 2018); (Houmani et al., 2018); (Dauwels et al., 2010); (Jeong, 2004). Electroencephalography (EEG), a non-invasive and cost-effective technique, stands as a promising modality to discern these changes (Thakor & Sherman, 2012); (Dauwels et al., 2010). EEG has proven to be invaluable, particularly in the detection and study of epilepsy, where it aids in detecting abnormal brain activity associated with seizures (Mansouri et al., 2012); (Smith, 2005). Previous studies have also explored the application of Graph Signal Processing (GSP) to model epileptic activity, showcasing the versatility and potential of utilizing EEG to detect clinically relevant conditions, for which we adapt to the context of dementia in this paper (Meena et al., 2022); (Mathur & Chakka, 2020).

## 2 Related Work

In recent years, the analysis of EEG signals has emerged as a promising avenue for the early detection of AD and several other neurodegenerative diseases (Cassani et al., 2018). Several studies have successfully harnessed the functional connectivity of electrode sites in EEG signals by feature engineering with GSP, utilizing standard machine learning (ML) models, or employing state-of-the-art Deep Learning (DL) methods such as Graph Neural Networks (GNNs) (Meena et al., 2022); (Mathur & Chakka, 2020); (Padole et al., 2018); (Song et al., 2021); (Vicchietti et al., 2023). This study contributes to this ongoing effort by benchmarking several ML models using GSP features created and the Graph Discrete Fourier Transform (GDFT) for AD detection in EEG data. We give an outline of the following sections below.

In (3.1) we provide a description of the dataset containing the EEG signals of AD, FTD, and Healthy Control (HC) patients. We describe our method for graph structure formation given from the raw EEG signal (3.2) and its derived GSP features including the GDFT (3.3). We benchmark these features on various ML and DL models (3.4), outlining model performance statistics in (4), before discussing our findings and conclusions in (5).

## 3 Methods

### 3.1 Data

We utilized the OpenNeuro ds004504 dataset, derived from recordings made at the 2nd Department of Neurology of AHEPA General Hospital, Thessaloniki (Miltiadous et al., 2023). The data was captured using a Nihon Kohden EEG 2100 clinical device fitted with 19 scalp and 2 reference electrodes, operating at a 500 Hz sampling rate and with a high-frequency filter at 70 Hz. The dataset contained three patient groups: AD, FTD, and HC, with average recording durations of 13.5 minutes, 12 minutes, and 13.8 minutes respectively. The cumulative durations for AD, FTD, and HC were 485.5 minutes, 276.5 minutes, and 402 minutes respectively.

During the preprocessing phase, raw data was transformed into a BIDS-compliant .set format, initially exporting it in .eeg format. The preprocessed and denoised recordings were stored in designated sub-0XX folders within a derivatives subfolder, segregating unprocessed and processed data. The preprocessing pipeline included a Butterworth band-pass filter at the 0.5-45 Hz range, re-referencing to A1-A2, and utilizing Artifact Subspace Reconstruction (ASR). Independent Component Analysis (ICA) was executed to shape the data into 19 ICA components, and automatic rejection of eye and jaw artifacts was implemented. The researchers abstained from including automatic annotations of artifacts to circumvent language compatibility issues. This preprocessed dataset can be found in the `derivatives` folder, containing the files used for this study. The following table below depicts additional information for each group including mean Mini-Mental State Examination (MMSE) and Median Disease Duration (MDD).

**Table 1:** Summary of patient characteristics for the OpenNeuro ds004504 dataset.

| Group | Characteristics |  |  |  |
| --- | --- | --- | --- | --- |
| | $n$ | Mean Age (years) | Mean MMSE | MDD (months) |
| Alzheimer Disease | 36 | $66.4 \pm 7.9$ | 25 | 13.5 |
| Frontotemporal Dementia | 23 | $63.6 \pm 8.2$ | N/A | 12 |
| Healthy Controls | 29 | $67.9 \pm 5.4$ | N/A | 13.8 |

**Figure 1:**
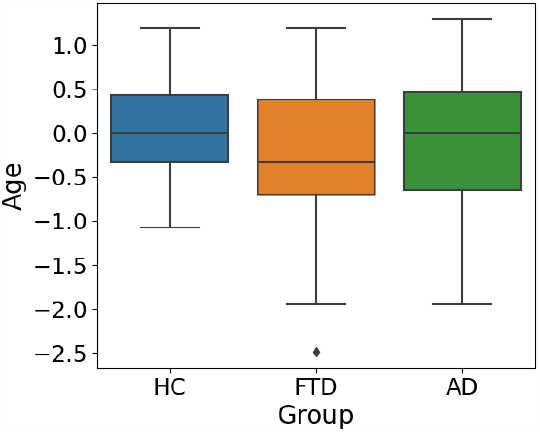
Box plot of age by group.

**Figure 2:**
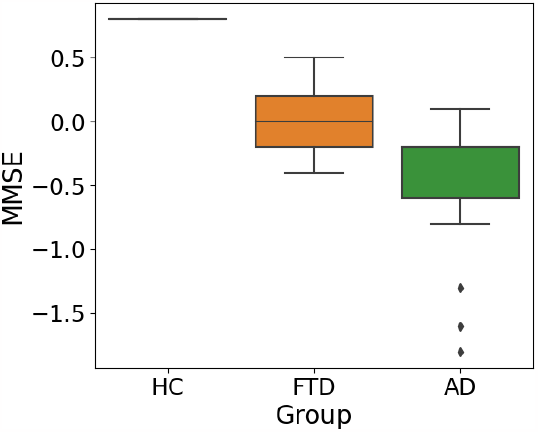
Box plot of MMSE by group.

### 3.2 Graph Structures

In graph theory, we define an undirected weighted graph by a triplet *𝒢* = (𝒱, ℰ, **W**), where 𝒱 is a set of nodes (or vertices) representing individual objects, and ℰ is an edge set representing relations between those objects with each element of the form (*i, j*, **W**^*i,j*^) for some *i, j* ∈ 𝒱 and **W** ∈ ℝ^| 𝒱 |*×*| 𝒱 |^ being the weight matrix that represents the quantitative relationship between each node pair. An EEG signal is a matrix **X** ∈ ℝ^*N×M*^ where *N* is the number of electrodes and *M* is the number of discrete time points, written as:

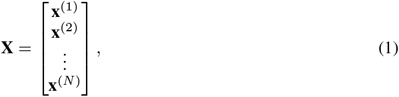

where each **x**^(*i*)^ ∈ ℝ^*M*^ is an individual electrode signal for 1 ≤ *i* ≤ *N*. Given **X**, a graph 𝒢 = (𝒱, ℰ, **W**) is formed where the vertex set 𝒱 = {1, 2 …, *N* } are the nodes representing the set of electrodes. The edge set ℰ and weight matrix **W** are created as follows. For parameters *θ, k* ∈ (0, ∞), we first define **W** ∈ ℝ^*N ×N*^ via a Gaussian kernel (ter Haar Romeny, 2003):

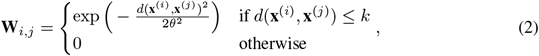

for each 1 ≤ *i, j* ≤ *N*, which represents the degree of connectivity between each electrode pair (*i, j*). For our experiments, we chose *d* to be the metric induced by the Euclidean norm ∥ *·* ∥_2_, i.e. *d*(**x, y**) = ∥ **x** − **y** ∥_2_. The edge set is defined as the indices of the nonzero entries in **W**:

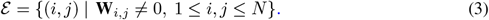

### 3.3 Graph Signal Processing

We introduce the relevant GSP methods to create the input features from the EEG signal **X** and graph 𝒢, before using these features to train our ML models in 3.4 (Ortega et al., 2018); (Dong et al., 2020);(Li et al., 2021). For standard methods in GSP see (Ortega, 2022). The first step is to obtain the **Combinatorial Graph Laplacian** matrix (Chung & Langlands, 1996):

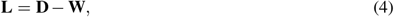

where **D** is the diagonal degree matrix defined by:

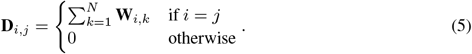

Since **L** is normal, by the Spectral Theorem we can write **L** = **U K U**^*^ where the columns of **U** ∈ ℝ^*N×N*^ are the eigenvectors of **L**. Here, **L** is a real symmetric matrix. Therefore, all eigenvalues and eigenvectors are real. Hence, we can simply write **L** = **U K U**^*T*^. The GDFT is given by (Mathur & Chakka, 2020):

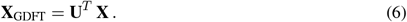

Let **C** be the covariance matrix of **X**_GDFT_. We project **C** onto the space of the Laplacian’s eigenvectors to obtain:

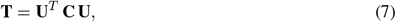

for which we compute the **Stationary Ratio** defined as:

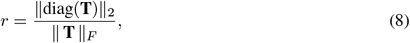

where ∥·∥_*F*_ denotes the Frobenius norm (Meena et al., 2022). Let **P** = **L X**. We define the **Tik-norm** by:

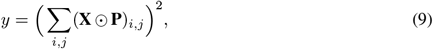

where ⊙ is the Hadamard product and the sum is taken over all entries of **X** ⊙ **P** (Meena et al., 2022). Other input features include:

1. **Total Variation (TV):** Total Variation measures the overall variation or dissimilarity in the data represented by matrix **X** (Sandryhaila & Moura, 2014). It is defined as:

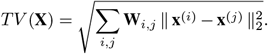

Higher Total Variation indicates greater dissimilarity among data points within the dataset.
2. **Graph Energy:** Graph Energy is a measure that captures the structural properties of a graph represented by the Laplacian matrix **L** (Balakrishnan, 2004). It is calculated by:

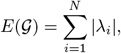

where *λ*_1_, …, *λ*_*n*_ are the eigenvalues of **L**. A higher Graph Energy indicates a more complex and interconnected graph structure.
3. **Spectral Entropy:** Spectral Entropy quantifies the uncertainty or randomness in the spectral distribution of a matrix **U** Rényi (1961). **It is defined as:**

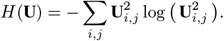

**Higher Spectral Entropy values indicate a more diverse and less predictable spectral distribution**.
4. **Signal Energy:** Signal Energy measures the total energy or magnitude of the data represented by matrix **X** (Rihaczek, 1968). It is computed as:

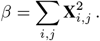

Higher Signal Energy values indicate data with higher overall magnitude of the signal.
5. **Signal Power:** Signal Power, denoted by *σ*^2^, is equivalent to the variance of the data in matrix **X** defined by:

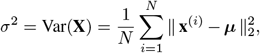

where 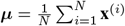 is the mean of the dataset **X** (Rao & Swamy, 2018). High Signal Power values indicate greater variability in the data.
6. **Unique Spectral Cluster Labels:** Unique Spectral Cluster Labels, denoted by *γ*, represents the number of distinct clusters or groups identified when applying spectral clustering to the weight matrix **W**. Spectral clustering is a technique for grouping data points based on their connectivity in a graph. *γ* indicates the number of distinct clusters found. In our experiments we applied the `spectral clutsering` method to **W** with the `scikit-learn` package to compute *γ* (Pedregosa et al., 2011).
7. **Average Degree:** Average Degree, denoted by *μ*_**D**_, quantifies the average connectivity of nodes in a graph represented by the degree matrix **D** (Ortega, 2022). It is calculated as:

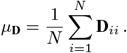

A higher *μ*_**D**_ indicates greater connectivity of the graph.
8. **Heat Trace:** Heat Trace, represented by *h*, is calculated by taking the trace (sum of diagonal elements) of a modified Laplacian matrix **L**^*′*^, where each element is exponentiated by 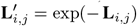 (Xiao et al., 2009). It is defined as:

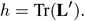

The Heat Trace is related to diffusion processes on graphs and can capture how information or “heat” spreads across the graph.
9. **Diffusion Distance:** Diffusion Distance, denoted by *h*^*′*^, measures the overall spread or extent of diffusion in a graph (Hammond et al., 2013). It is calculated with:

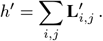

The Diffusion Distance of a graph provides insight into how information or influence propagates across the graph.

For each patient EEG signal, we generate the following features using the methods above:

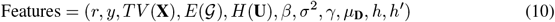

**Figure 3:**
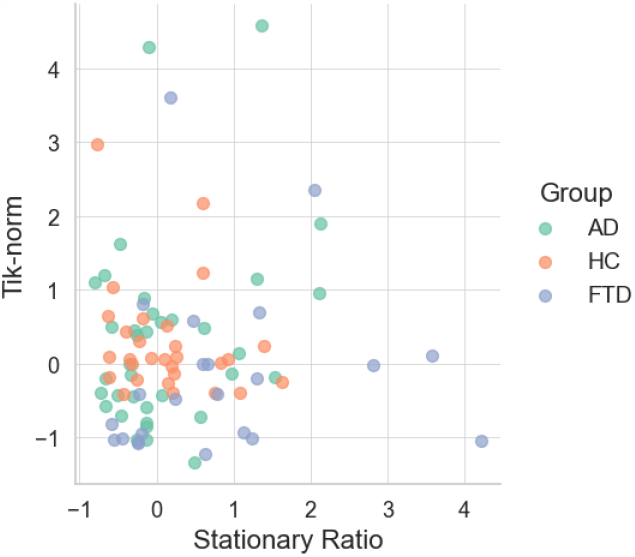
Plot of stationary ratio versus Tik-norm by group for AD, FTD, and HC.

**Figure 4:**
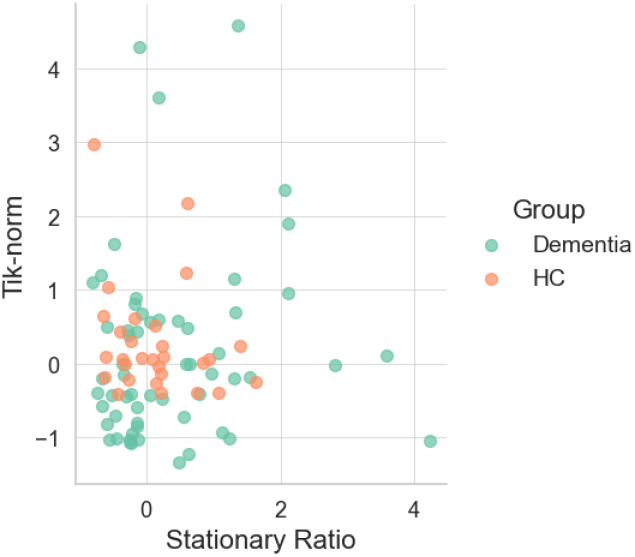
Plot of stationary ratio versus Tik-norm by group for Dementia and HC.

**Figure 5:**
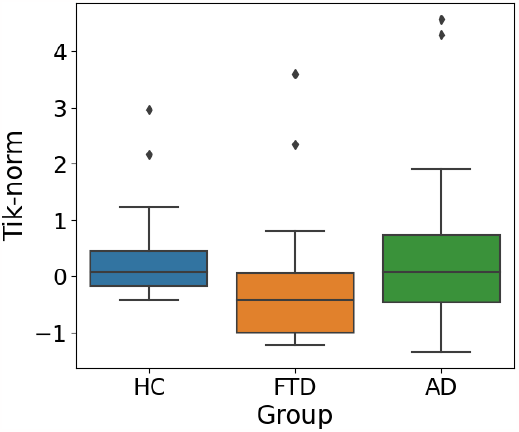
Box plot of Tik-norm by group.

**Figure 6:**
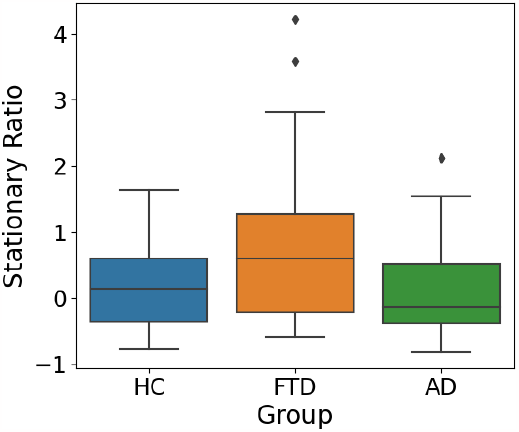
Box plot of stationary ratio by group.

**Figure 7:**
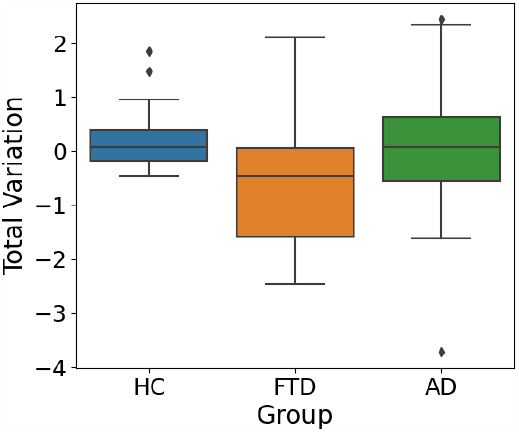
Box plot of total variation by group.

**Figure 8:**
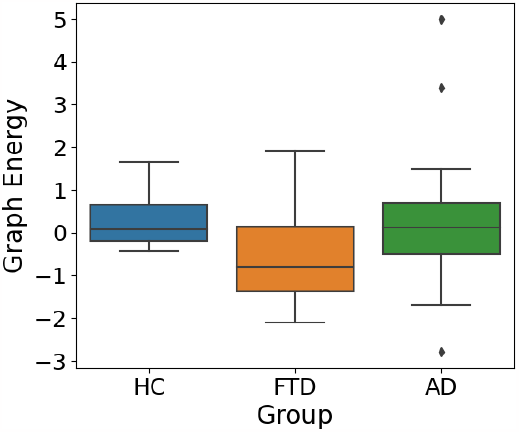
Box plot of graph energy by group.

**Figure 9:**
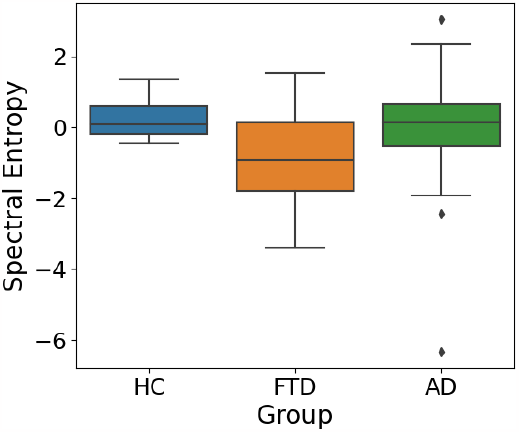
Box plot of spectral entropy by group.

**Figure 10:**
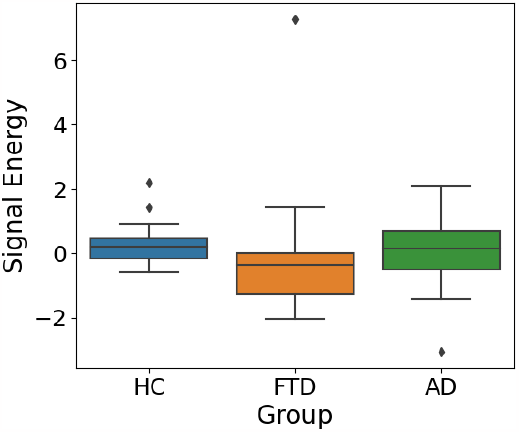
Box plot of signal energy by group.

**Figure 11:**
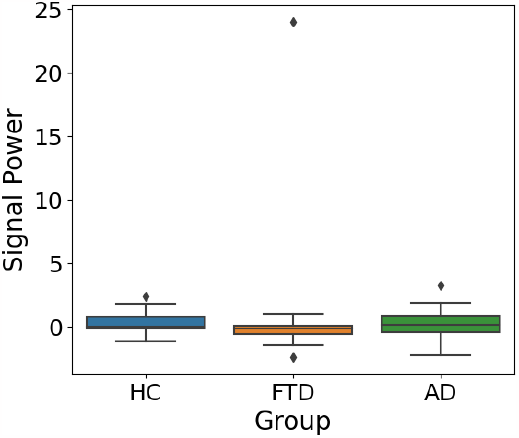
Box plot of signal power by group.

**Figure 12:**
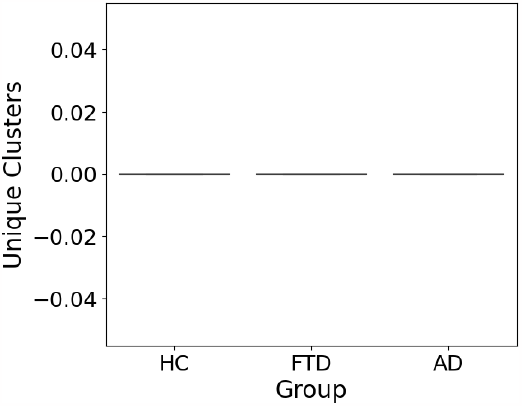
Box plot of unique spectral clusters by group.

**Figure 13:**
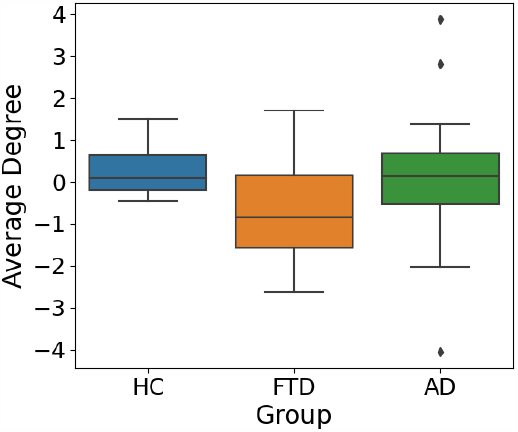
Box plot of average degree by group.

**Figure 14:**
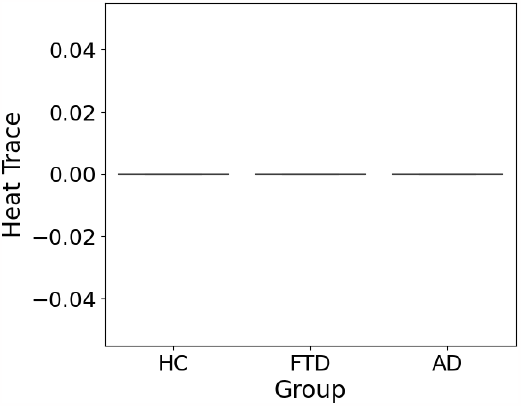
Box plot of heat trace by group.

**Figure 15:**
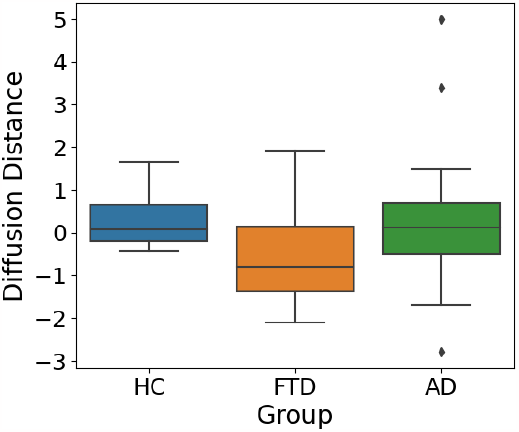
Box plot of diffusion distance by group.

**Figure 16:**
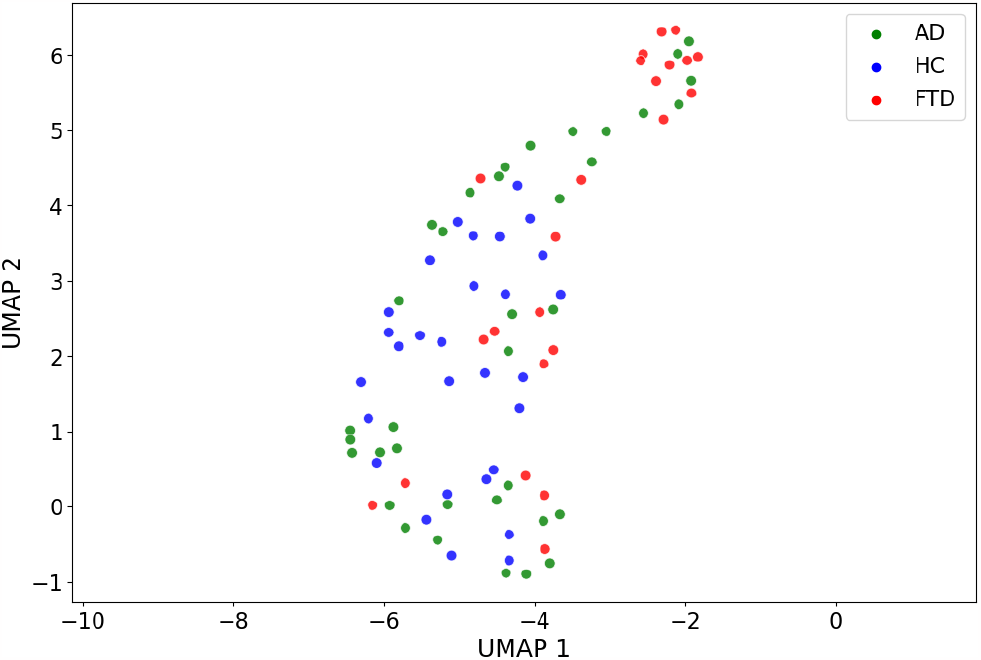
Plot of UMAP projected EEG features by group.

### 3.4 Models

We test our features from Equation 10 by training and evaluating several ML models below on the task of multiclass classification (AD, FTD, HC), and binary classification (Dementia, HC) where the Dementia group comprises of AD and FTD.

a. *k***-Nearest Neighbours (***k***-NN) Classification** : Given a new observation **x**_0_, the *k*-NN algorithm searches through the training dataset to find the *k* training examples that are closest to **x**_0_ based on a distance metric (e.g., Euclidean distance). The predicted label is then obtained by majority voting among the *k* nearest neighbours (Fix & Hodges, 1989).
b. **Random Forest**: A Random Forest is a collection of decision trees {*h*(**x**; Θ_*k*_), *k* = 1, …}, where Θ_*k*_ are the parameters of each tree. A decision tree is a flowchart-like structure where each internal node represents a feature (or attribute), the branch represents a decision rule, and each leaf node represents the outcome. The root node is the feature that best splits the dataset into classes based on a decision criterion, and this process is recursively applied to the sub-sets of data reaching each internal node, constructing the tree until it reaches a specified depth or purity. The final prediction in a Random Forest is obtained by averaging the predictions of all the trees for regression, or by majority voting for classification (Ho, 1995).
c. **XGBoost**: XGBoost builds an ensemble of decision trees. The prediction is given by 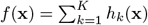, where *h*_*k*_(**x**) is an individual tree, and *K* is the number of trees (Chen & Guestrin, 2016).
d. **Logistic Regression**: Logistic Regression is a statistical model used for binary classification, where the probability of the positive class is modeled as 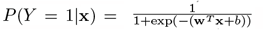, where **w** is the weight vector, and *b* is the bias term (Menard, 2002). To convert this probability into a binary outcome, a threshold value, typically 0.5, is chosen. If *P* (*Y* = 1|**x**) ≥ 0.5, the prediction is 1; otherwise, the prediction is 0. The threshold can be adjusted based on the specific needs of a problem, such as minimizing false positives or negatives. In a multiclass setting, Logistic Regression can be extended using techniques like One-Versus-Rest (OvR) or Multinomial Logistic Regression, where the model estimates the probability of each class and picks the class with the highest estimated probability.
e. **Support Vector Machine (SVM)**: SVM finds the hyperplane that maximizes the margin between the two classes. The decision function is given by *f* (**x**) = **w**^*T*^ **x**+*b*, where **w** is the normal vector to the hyperplane (a subspace of 1 dimension lower) and *b* is the bias term (Cortes & Vapnik, 1995). For instance, in a 2-dimensional space (i.e. the Cartesian plane), a hyperplane is simply a line. If we have two classes of points scattered on the plane, SVM will find the line that best separates these two classes while keeping the maximum distance from the nearest points of each class to this line.
f. **Naive Bayes**: Naive Bayes is a probabilistic classifier based on applying Bayes’ Rule, which relates the conditional and marginal probabilities of random events, with the “naive” assumption of independence between features. Bayes’ Rule is expressed as 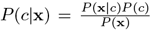. However, computing *P*(**x**|*c*) directly can be challenging due to the high dimensionality of **x**. The naive independence assumption simplifies this to 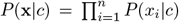, leading to the formula 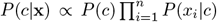, where *n* is the number of features (Rish et al., 2001). This simplification allows for efficient computation and training of the model. The classifier then predicts the class *c* that maximizes *P* (*c*|**x**), i.e., the class that is most probable given the feature vector **x**.
g. **TabTransformer**: TabTransformer employs self-attention mechanisms to capture interactions among features in tabular data (Huang et al., 2020). The self-attention mechanism is a key component of the transformer architecture, which allows the model to weigh the importance of different parts of the input differently when making predictions. Specifically, self-attention computes a weighted sum of all input features, where the weights are determined by the similarity between the query and key representations of the features, allowing each feature to attend over all others in a flexible manner. The self-attention formula is given by Attention 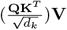, where **Q, K**, and **V** are the query, key, and value matrices, respectively, and *d*_*k*_ is the dimension of the key vectors (Vaswani et al., 2017). This mechanism enables TabTransformer to effectively capture feature interactions in tabular data, even when relationships among features are complex and non-linear.

## 4 Results

For each model, we utilized the full dataset, splitting it into a training set (35%), validation set (35%), and test set (30%). Tables 2 and 3 display accuracy for each classification task and model, and Tables 4 and 5 display precision, recall, F1-score, and support for each class and each task.

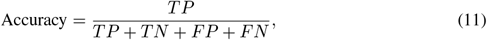

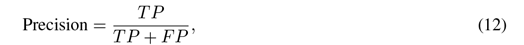

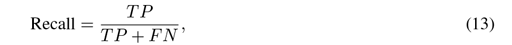

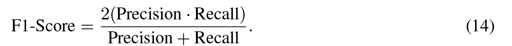

**Table 2:** Model performance on the test set for multiclass classification of AD, FTD, and HC. Highest accuracy is bolded.

| Method | Accuracy |
| --- | --- |
| $k$-NN Classifier | 0.37 |
| Random Forest | 0.33 |
| XGBoost | 0.41 |
| Logistic Regression | 0.41 |
| SVM | 0.37 |
| Naive Bayes | **0.44** |
| TabTransformer | 0.21 |

**Table 3:** Model performance on the test set for binary classification of dementia (AD, FTD) versus HC. Highest accuracy is bolded.

| Method | Accuracy |
| --- | --- |
| $k$-NN Classifier | 0.70 |
| Random Forest | 0.67 |
| XGBoost | 0.74 |
| Logistic Regression | 0.63 |
| SVM | **0.85** |
| Naive Bayes | 0.59 |
| TabTransformer | 0.71 |

**Table 4:** Other model performance metrics on the test set for multiclass classification of AD, FTD, and HC.

| Method | Precision | Recall | F1-Score | Support |
| --- | --- | --- | --- | --- |
| **AD Classification** |  |  |  |  |
| $k$-NN Classifier | 0.50 | **0.54** | **0.52** | 13 |
| Random Forest | 0.40 | 0.46 | 0.43 | 13 |
| XGBoost | 0.44 | 0.31 | 0.36 | 13 |
| Logistic Regression | **0.67** | 0.31 | 0.42 | 13 |
| SVM | 0.60 | 0.23 | 0.33 | 13 |
| Naive Bayes | 0.50 | 0.15 | 0.24 | 13 |
| TabTransformer | 0.25 | 0.14 | 0.18 | 7 |
| **FTD Classification** |  |  |  |  |
| $k$-NN Classifier | **1.00** | 0.20 | 0.33 | 10 |
| Random Forest | 0.00 | 0.00 | 0.00 | 10 |
| XGBoost | 0.67 | 0.40 | 0.50 | 10 |
| Logistic Regression | 0.75 | 0.30 | 0.43 | 10 |
| SVM | **1.00** | 0.30 | 0.46 | 10 |
| Naive Bayes | 0.75 | **0.60** | **0.67** | 10 |
| TabTransformer | 0.50 | 0.17 | 0.25 | 6 |
| **HC Classification** |  |  |  |  |
| $k$-NN Classifier | 0.09 | 0.25 | 0.13 | 4 |
| Random Forest | 0.25 | 0.75 | 0.38 | 4 |
| XGBoost | 0.25 | 0.75 | 0.38 | 4 |
| Logistic Regression | 0.24 | **1.00** | 0.38 | 4 |
| SVM | 0.21 | **1.00** | 0.35 | 4 |
| Naive Bayes | **0.27** | **1.00** | **0.42** | 4 |
| TabTransformer | 0.12 | **1.00** | 0.22 | 1 |

**Table 5:**
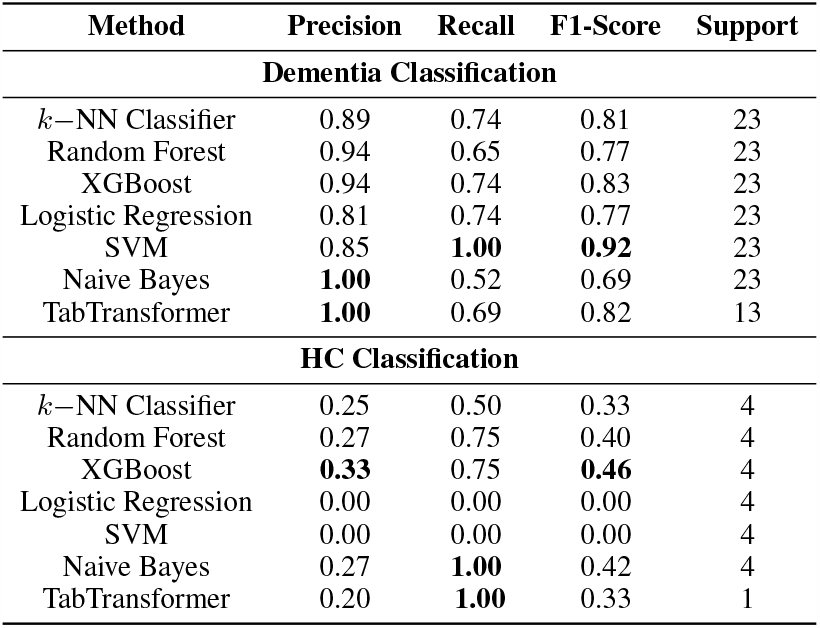
Other model performance metrics on the test set for binary classification of Dementia and HC.

Here, *T P, T N, FP*, and *FN* are the number of true positives, true negatives, false positives, and false negatives respectively. Precision measures the reliability of a model’s positive classifications, while recall gauges its completeness in capturing positive samples. The F1-score offers a balanced evaluation of both, using their harmonic mean to provide a holistic view of model performance. Support refers to the number of occurrences of each class.

Table 2 displays Naive Bayes as the model with highest test accuracy at 44%, while the TabTransformer model had the lowest accuracy, achieving 21%. Aside from the TabTransformer, model performance is relatively close, implying that the TabTransformer may be overfitted to the training dataset, with poor generalization to the test set. With the strong assumption of independence of the features in Naive Bayes in 3.4, it may be the case that the features from 10 are sufficiently independent to harness the advantage of Naive Bayes. However, with a top accuracy that is just above random guessing rate (33%), this suggests that the choice of model is likely not to be substantially impacting accuracy, and points to an issue of separability of the classes and the inherent data distribution for AD, FTD, and HC; or the inability for the features in Equation (10) to incorporate enough information to distinguish between the three classes.

Table 3 presents the performance of each model in the binary classification setting, trained to distinguishing between cases of dementia (AD, FTD) and HC. The SVM model, employing a linear kernel, displayed the highest accuracy of 85%, while the Naive Bayes model lagged with an accuracy of 59%. The SVM model’s superior performance suggests that the dataset may be largely linearly separable, although this should be further validated (Cortes & Vapnik, 1995). Conversely, the suboptimal performance of Naive Bayes is likely attributable to the violation of its feature independence assumption, signifying that the features are likely dependent in this binary classification context (Rish et al., 2001).

Table 4 further evaluates each models’ performance in the multiclass classification setting, displaying precision, recall, F1-Score, and support. For AD classification, Logistic Regression shows the highest precision at 67%, making it the most reliable in terms of positive predictive value, possibly due to its parametric nature which might be capturing the underlying data distribution effectively (Menard, 2002).

In FTD classification, *k* − NN and SVM both achieve perfect precision but differ substantially in recall and F1-Score; this could be indicative of *k*-NN’s sensitivity to noise and outliers, as well as SVM’s capacity for finding a more discriminative hyperplane in the feature space (Fix & Hodges, 1989); (Rish et al., 2001). Naive Bayes emerges as the most balanced model for FTD with the highest F1-Score of 67%, perhaps owing to its probabilistic framework that may better capture the conditional dependencies among features (Rish et al., 2001).

For HC, all models exhibit low precision, with Naive Bayes performing the best at 27%. This may indicate that the feature representation is not robust enough for effectively distinguishing this class, suggesting that the current features in Equation 10 are not sufficient for the task of multiclass classification. Additionally, given a low support of only *n* = 4 for HC, as a function of the relatively small dataset size of *n* = 88, values within the HC table are highly susceptible to the outcome of this random sample.

Table 4 extends the evaluation to the binary classification task, assessing the models’ ability to distinguish between dementia cases and HC. In the dementia setting, all models scored relatively high for precision. Although, Naive Bayes achieved perfect precision, it had the lowest recall of 52% suggesting that its assumptions of feature independence may only hold in the positive class Rish et al. (2001). SVM achieved relatively high precision and perfect recall, suggesting the data distribution is likely linear separable in these two classes Cortes & Vapnik (1995). All models achieved relatively high F1-score with SVM being the highest.

The precision for all models in the HC classification was notably lower compared to the dementia classification, suggesting that while our features from Equation (10) are effective in identifying dementia traits, they may not be sufficient to distinguish attributes of healthy individuals. It is also worth noting that due the support value of 4, the values in HC Classification section are potentially subject to fluctuations and can vary by training run.

## 5 Conclusion

Our method introduces a novel approach utilizing GSP techniques with the GDFT for generating features to detect AD in EEG recordings. SVM was accurate in differentiating between dementia and HC in 85% of cases. However distinguishing AD from FTD was challenging in the multiclass classification setting across all models, yielding a top accuracy of only %44. We conclude that our features from Equation (10) are useful for identifying healthy versus dementia patients, but they are not applicable for determining the type of dementia a patient might have.

The discrepancy between performance in binary classification and multiclass classification suggests that distinguishing AD and FTD is significantly more challenging and may require a more nuanced approach. We speculate that this challenge arises due to: (1) the similarities between AD and FTD features given by GSP; (2) the wide variability in FTD types (e.g., sporadic, behavioural); (3) the variability of disease stages in AD; (4) the lack of model fit (e.g., TabTransformer overfitting, traditional ML models underfitting).

In future work, we aim to address the variation seen within AD by incorporating multiclass classification of different stages of AD, including the pre-clinical, mild, moderate and severe stages, as well as implementing a similar approach for FTD types. This would not only provide more information for the machine learning model to use, but would also allow for potential earlier disease detection and informed diagnoses to prevent subsequent worsening of symptoms. We acknowledge the limitations of using mostly standard machine learning models, so we plan to implement state-of-the-art deep learning (DL) methods such as graph neural networks (GNNs) that utilize node features, edge features, and graph-level features given by our GSP method. It is worth noting that these methods are useful to explore but require technical proficiency to integrate into a clinical setting. Therefore, future work can also include clinician education and improving ease of use.

## Data Availability

The original dataset used in the study can be found at the original link here: https://openneuro.org/datasets/ds004504/versions/1.0.6
Code for generating features, plots, and training machine learning models for reproducibility can be found here: https://github.com/xmootoo/gsp-alzheimer-detection

## 6 Acknowledgements

We would like to express our gratitude to the researchers from the Department of Informatics and Telecommunications at the University of Ioannina, the Department of Electrical and Computer Engineering at the University of Western Macedonia, and the 2nd Department of Neurology at AHEPA University Hospital, Aristotle University of Thessaloniki, for generously sharing their dataset Miltiadous et al. (2023). Their open collaboration greatly contributed to the advancement of our research.

Xavier Mootoo also gratefully acknowledges the support received from the Vector Institute through the Vector Scholarship in AI and the Natural Sciences and Engineering Research Council of Canada (NSERC) through the Canada Graduate Scholarship—Master’s.

